# *PIEZO1* variants that reduce open channel probability are associated with familial osteoarthritis

**DOI:** 10.1101/2024.09.03.24312969

**Authors:** Michael J. Jurynec, Elena Nosyreva, David Thompson, Crystal Munoz, Kendra A. Novak, Derek J. Matheson, Nikolas H. Kazmers, Ruhma Syeda

## Abstract

The synovial joints senses and responds to a multitude of physical forces to maintain joint homeostasis. Disruption of joint homeostasis results in development of osteoarthritis (OA), a disease characterized by loss of joint space, degeneration of articular cartilage, remodeling of bone and other joint tissues, low-grade inflammation, and pain. How changes in mechanosensing in the joint contribute to OA susceptibility remains elusive. PIEZO1 is a major mechanosensitive cation channel in the joint directly regulated by mechanical stimulus. To test whether altered PIEZO1 channel activity causes increased OA susceptibility, we determined whether variants affecting *PIEZO1* are associated with dominant inheritance of age-associated familial OA. We identified four rare coding variants affecting *PIEZO1* that are associated with familial hand OA. Single channel analyses demonstrated that all four PIEZO1 mutant channels act in a dominant-negative manner to reduce the open probability of the channel in response to pressure. Furthermore, we show that a GWAS mutation in *PIEZO1* associated with reduced joint replacement results in increased channel activity when compared with WT and the mutants. Our data support the hypothesis that reduced PIEZO1 activity confers susceptibility to age-associated OA whereas increased PIEZO1 activity may be associated with reduced OA susceptibility.

## Introduction

The synovial joint is a mechanosensitive organ that responds to various physical forces including compressive, shear, and hydrostatic stresses^1,2^. These joint cells sense (mechanosensing) and respond (mechanotransduction) to daily physical forces to maintain homeostasis of the joint. Disruption of mechanosensing or mechanotransduction can lead to molecular and cellular changes in the joint that are associated with the development of osteoarthritis (OA)^3–7^, which is characterized by loss of joint space, degeneration of articular cartilage, remodeling of bone and other joint tissues, low-grade inflammation, and pain^8,9^. Many physiological and environmental risk factors are associated with increased risk of developing OA, including genetics, traumatic joint injury, aging, obesity, and altered biomechanics. Mechanosensing and mechanotransduction can be influenced by many of the same factors, which can be exacerbated by mechanical overloading, injury or unloading^10–14^. Despite the importance of mechanobiology in joint homeostasis and disease, we do not fully understand how alterations of the proteins that sense and respond to physical forces contribute to the development of OA.

PIEZO1 and TRPV4 are the key mechanosensitive cation channels in the joint directly regulated by mechanical stimulus. Previous *in vitro* studies using chondrocytes (the major cell type of cartilage) have indicated that PIEZO1 is activated at high levels of cellular deformation (e.g. hyperphysiological load such as a traumatic injury) and TRPV4 is activated under normal physiological load^15–19^. Upon activation there is an influx of Ca^2+^ and Na^+^ into the cell which leads to cell volume changes, activation of signaling pathways and changes in gene expression^4,5,16,17,20,21^. Activation of PIEZO1 in the joint has been proposed to promote the OA phenotype by inducing inflammatory and catabolic gene expression, cell death, and senescence^3,15,16,21,22^. In line with this, i*n vivo* studies using a nonspecific PIEZO1 antagonist (GsMTx4) suggests that inhibition of PIEZO1 is protective against injury-induced OA in rodent models^23,24^. In contrast to PIEZO1, TRPV4 activation in the joint is largely considered necessary to maintain homeostasis of the joint through activation of anabolic gene expression^19,21,25,26^. While previous studies have implicated hyperphysiological activation of PIEZO1 to elicit expression of OA-associated markers, a recent study has also demonstrated that activation of PIEZO1 with a chemical agonist promotes expression of pro-chondrogenic genes and increased deposition of sulfated glycosaminoglycans^21^. These data suggest that PIEZO1 may have context dependent roles in maintaining homeostasis of the joint.

Despite the significant genetic contribution to OA^27–31^, there has only been one reported genetic association of *PIEZO1* variants with the OA phenotype^32^. A genome wide association study (GWAS) identified a dominant rare coding allele of *PIEZO1* that was associated with reduced OA progression^32^. While this allele was associated with reduced OA progression (as defined by joint replacement), it is unknown how this coding mutation affects PIEZO1 channel activity. No molecular studies were performed to determine if this mutation alters PIEZO1 channel activity. Therefore, we cannot determine if reduced or increased PIEZO1 activity is associated with reduced OA progression in humans. In sum, the published data indicates that the role of PIEZO1 in OA is not resolved and that the channel may have context dependent or tissue specific roles in acute injury vs aging.

The genetic analysis of families with dominant forms of OA allows us to identify highly penetrant coding alleles that have determinate effect promoting OA, independent of prior biases on how protein activity may affect the OA phenotype or the tissues specific requirement of the mutant alleles^33–40^. Non-null alleles further allow for functional studies to determine how the coding mutations affect protein activity and function. For example, compound heterozygous coding variants of *PIEZO1* were discovered in an individual with Prune Belly Syndrome. Extensive single channel functional analyses demonstrated the PIEZO1 mutations reduced the pressure-induced open probability of the channel^41^. Here we report the identification of four families with age-associated OA with independent OA-susceptibility alleles in *PIEZO1*. All four *PIEZO1* mutant channels have reduced activity, while the GWAS mutation (F2484L) results in increased channel activity when compared with WT. Our data indicate that reduced PIEZO1 activity increases susceptibility to age-associated OA and increased PIEZO1 activity may be protective in the absence of acute injury. We hypothesize that PIEZO1 may have context dependent or tissue specific roles in injury vs aging.

## Results

### Identification of *PIEZO1* Variants in Families with Age-Associated OA

We took an unbiased genetic approach to determine if mutations in *PIEZO1* have a strong effect on OA susceptibility by utilizing a large statewide medical genetics database, the Utah Population Database^33,34,36,42^. We identified a cohort of 151 families with dominant forms of non-syndromic familial OA that were free from acute or traumatic joint injury^33–36^. Each family is characterized by a defining form of OA that affects a distinct subset of joints, including erosive hand OA (EHOA)^36,43,44^ and interphalangeal (IP) joint OA^45,46^. Although the families are identified by shared OA phenotype, individuals in a family often have OA in additional joints, including the large weight bearing joints such as the hip, knee, and spine **(Table 1).**

**Table 1.**
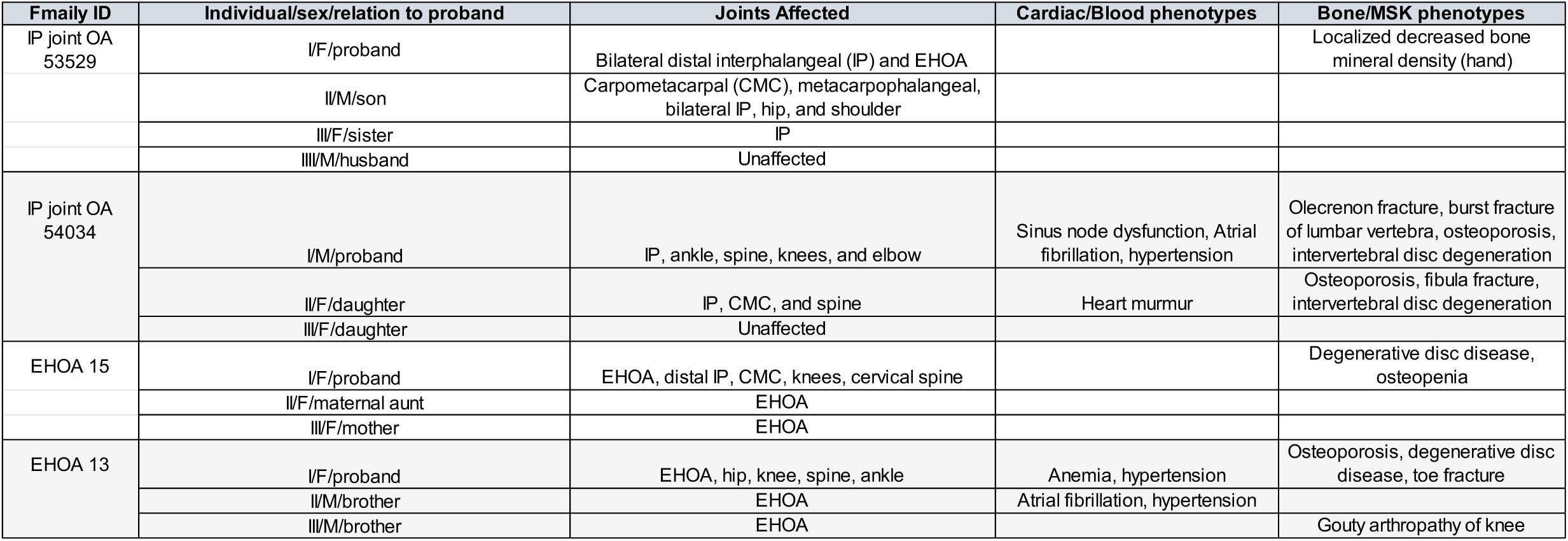
Osteoarthritis Families and Phenotype Details.

We performed whole exome sequence analysis on informative members (selected affected and unaffected individuals) of families and identified coding variants that invariably segregated with OA^34,35^. We identified rare *PIEZO1* coding variants in four independent families diagnosed with bilateral EHOA or IP joint OA **(Figure 1a, 1b and Tables 1 and 2)**. Among the analyzed family members, *PIEZO1* variants were carried by all individuals with OA and absent from disease-free individuals. All variants were evolutionarily conserved in vertebrates, rare in the general population (minor allele frequency < 0.01) and predicted to be damaging or disease causing.

**Figure 1.**
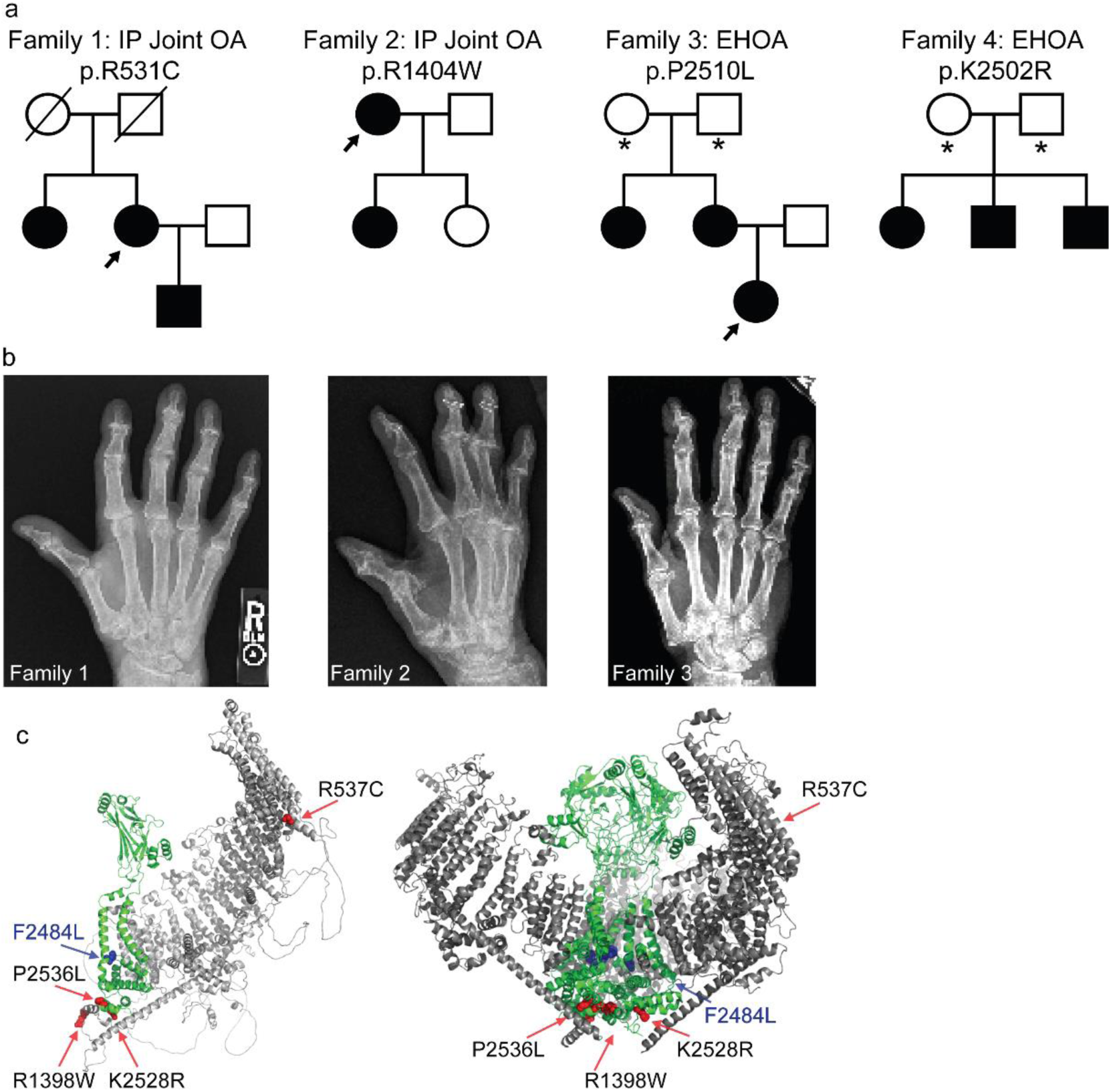
Dominant mutations in PIEZO1 associate with hand OA. **(a)** Interphalangeal (IP) joint OA and erosive hand OA (EHOA) segregate as apparent dominant traits in four independent families. The PIEZO1 mutation identified in each family is noted above the pedigree. Circles = females, squares = males, slash = deceased. Filled circles/squares = affected individuals; open circles/squares = individuals with no known history of hand OA. Asterisks = individuals with an unknown hand OA diagnosis. **(b)** Hand radiographs of individuals marked with an arrow in the high-risk pedigrees. **(c)** AlphaFold2 PIEZO1 predicted monomer structure (left) and Cryo-EM based mouse PIEZO1 trimeric structure (PDB ID #6B3R) (right). PIEZO1 blades (grey) encompass residues 1 to 2109. Pore domain (green) spans from residues 2110 to 2547. Familial mutations (mouse amino acid numbers) are shown as red spheres, GWAS mutation as blue spheres on a PIEZO1 monomer. Human mutations and corresponding mouse amino acids in parenthesis: p.R531C (p.R537), p.R1404W (p.R1398), p.K2502R (p.K2528), p.P2510L (p.P2536), p.F2458L (p.F2484L).

**Table 2.** PIEZO1 Variants Identified in Independent Osteoarthritis Families.

| OA Phenotype | Human Variant (Mouse Amino Acid) | rsID/Minor Allele Frequency | PolyPhen-2 and MutationTaster/CADD score |
| --- | --- | --- | --- |
| IP joint OA 53529 | p.R531C (p.R537) | rs146505418/0.0008618 | Damaging/32.0 |
| IP joint OA 54034 | p.R1404W (p.R1398) | rs188966111/0.0006523 | Damaging/32.0 |
| EHOA 15 | p.P2510L (p.2536) | rs61745086/0.00839 | Damaging/27.5 |
| EHOA 13 | p.K2502R (p.K2528) | rs34830861/0.005809 | Damaging/27.5 |

When mapped onto the mouse PIEZO1 Cryo-EM structure the mutations are in key functional regions of the protein. Two mutations, p.K2502R and p.P2510L, are in the PIEZO1 pore domain, and mutation p.R531C is in the peripheral blade region and is predicted to be near the inner leaflet of the lipid bilayer. The p.R1404W mutation is in a structurally unresolved region just before the beam that converges near the intracellular mouth of the channel and in proximity to the mutations in the pore **(Fig 1c).** Given the importance of PIEZO1 function in multiple tissues and organ systems^5^ and the association of *PIEZO1* mutations with hematological phenotypes^47–49^ and bladder defects^41^, we analyzed the OA families for additional phenotypes. While none of the phenotypes were completely penetrant, osteopenia and osteoporosis were present in some individuals in all four families and several cardiac defects, including hypertension, were noted in individuals in two of the families **(Table 2).** Our genetic studies indicate a striking correlation between inheritance of variants in *PIEZO1* and the occurrence of disease within families exhibiting EHOA and IP joint OA.

### Single Channel Analysis of OA-Associated PIEZO1 Variants

The OA-associated PIEZO1 mutations are predicted to be damaging, but *in silico* predictions fail to determine if the mutations are gain- or loss-of-function. Furthermore, both gain- and loss-of-function PIEZO1 mutations are associated with human disease phenotypes^5^. Therefore, it is important to perform functional analysis to assess whether the mutations are causal and alter normal PIEZO1 function. We generated expression vectors encoding the mouse WT and five mutant PIEZO1 channels (see Materials and Methods). The mouse amino acids corresponding to the human mutations are indicated in **Table 2**. We expressed these constructs in HEK293T^ΔP1^ cells that are null for endogenous PIEZO1 to obtain and compare WT and mutant channel function. In this context, normal WT PIEZO1 function refers to biophysical properties, such as permeation (single channel currents) and gating (open probability in response to applied pressure) properties obtained in cells. All the mutants yielded single channel activity in cell-attached mode **(fig 2 a),** with no significant difference in single channel currents obtained from all point current amplitude histograms **(Fig. 2b).** As expected for mechanosensitive channel, the open probability and channel activity increases when direct pressure is applied to the patch of membrane expressing WT, familial mutants R537C and P2536L, and GWAS F2484L mutant. The fold increase in the open probability after applied pressure was WT = 4.6, R537C = 3.5, P2536L = 2.2, F2484L = 3.2-fold **(Fig. 2c).** However, no significant difference was found before and after pressure application for the R1398W and K2528R, suggesting that the mechanosensitive gating properties are severely impaired in these mutations **(Fig. 2c).** Overall comparison of WT single channel properties with OA associated mutants clearly indicate no significant difference in conductance **(Fig. 2d),** while sharp decrease in open probability of familial mutants to that of WT PIEZO1 when assayed at -30 mm Hg pressure **(Fig. 2e).** In contrast, F2484L open probability was significantly higher than that of WT. These data indicate that the four familial PIEZO1 mutations are hypomorphic (loss-of-function), whereas the GWAS mutation is hypermorphic (gain-of-function).

**Figure 2.**
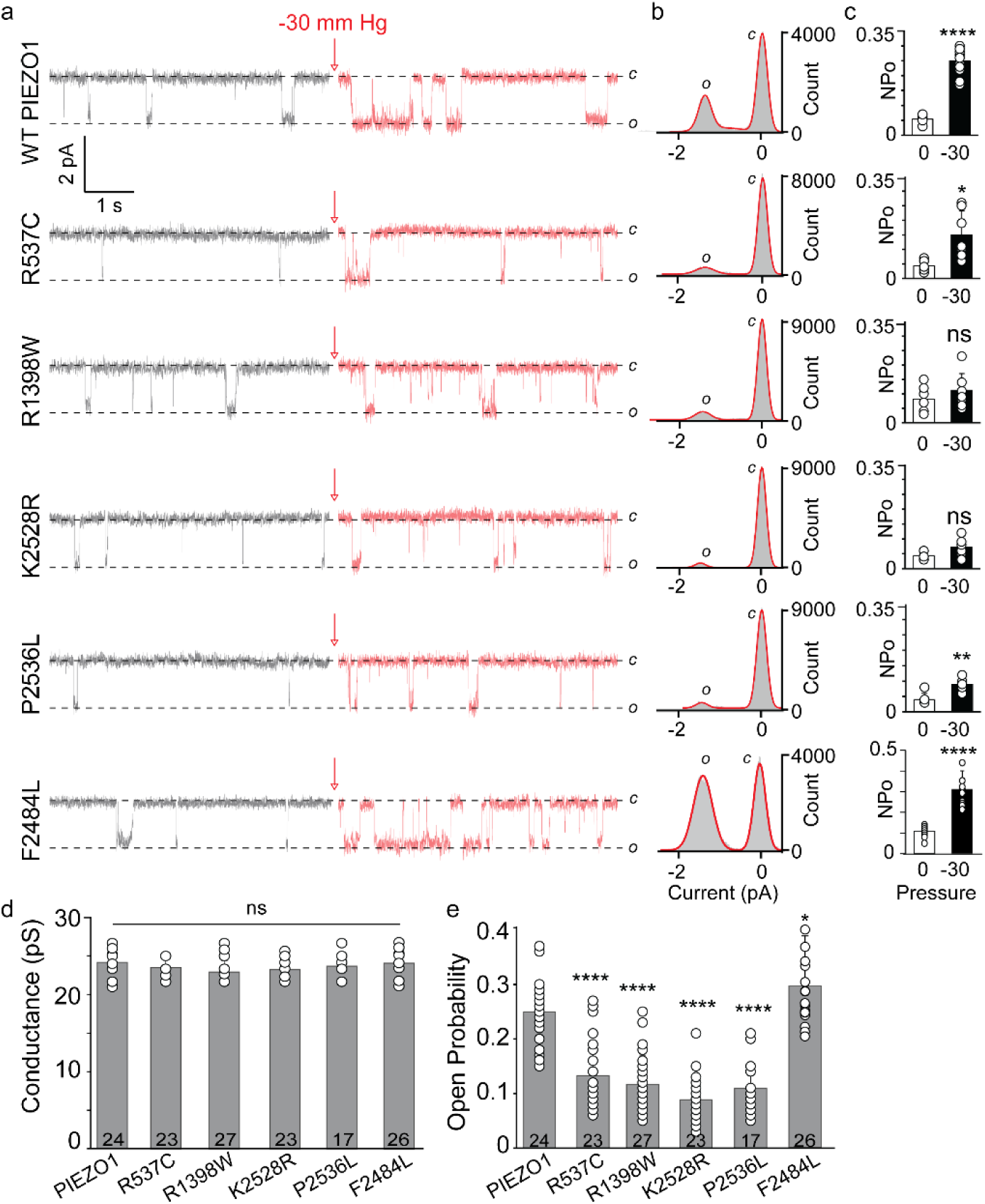
Single-channel properties of WT and OA-associated PIEZO1 channels obtained from cell-attached patch clamp recordings. **(a)** HEK293T^ΔP1^ were transfected with WT or mutant constructs. Single-channel current recordings of WT and mutant PIEZO1 proteins R537C, R1398W, K2528R, P2536L, and F2484L showing the closed (c) ∼0 pA and open (o) ∼-1.5 pA state of the channel. The pipette solution contained 130 mM Na+ and 1 mM Ca++, the bath solution contained 140 mM K+ and 1 mM Mg++. Red arrow and trace indicate application of -30 mm Hg pressure. **(b)** All-point current amplitude histograms of the single-channel recordings shown in panel a. The area under the curve demonstrates the c and o states of the channel. X-axis is the current amplitude in picoamperes (pA). Y-axis is total counts. **(c)** Steady state open channel probability of WT and mutant PIEZO1 channels with (black bars) and without (white bars) applied pressure (-30 mm Hg) to the patch of membrane. X-axis is pressure in mm Hg. Y-axis is Normalized Open Probability (NPo). **(d)** Single channel conductance of WT and mutant PIEZO1 channels obtained from panel b. Experimental replica numbers are shown in the bar graph (n>16). **(e)** Steady state open probability of WT and mutant PIEZO1 channels obtained from the area under the curve from the open state. ****p<0.0001 and ns = not significant.

All the familial OA-associated *PIEZO1* alleles segregate dominantly with the OA phenotype. Affected individuals have one WT allele and one mutant allele. To recapitulate these pathophysiology relevant conditions in the cellular *in vitro* assays, we co-expressed WT PIEZO1 (fused with TdTomato) and the mutants (fused with GFP) in HEK293T^ΔP1^ cells. Only the cells exhibiting both red and green signals were selected for patch clamp recordings to ensure the expression of both the WT PIEZO1 (red) and OA-associated familial variants (green). Structural studies have concluded that PIEZO1 is a homo-trimeric channel where three subunits assemble to form a central ion conduction pore. The co-expression of two PIEZO1 constructs can potentially produce three major combinations of PIEZO1 channels; i) Homo-trimers of WT PIEZO1, ii) homo-trimers of familial mutant channels, and iii) hetero-trimers of WT and mutant subunits (either in 2:1 or 1:2 subunit ratios). Since WT and familial mutants (when expressed alone) exhibit significantly different single-channel open probability **(Fig. 2e**), we relied on pressure-dependent gating properties to assess whether co-expression produces more prevalent WT-like activity or mutant-like activity. Surprisingly, co-expression of WT PIEZO1 and mutant produces activity that closely resembles familial mutant’s open probability **(Fig 3a)** and significantly different from WT (p<0.0001) **(Fig 3b).** Whereas no significant difference was found when the co-expression conditions were compared to the mutants expressed alone (p>0.095), suggesting a functional dominant negative effect of these mutations in HEK293T^ΔP1^ cells **(Fig 3).** As expected from data obtained in single mutant expression, no significant difference was found in single channel current amplitude between different combinations of PIEZO1 and familial mutant expressions, suggesting that permeation properties were intact.

**Figure 3.**
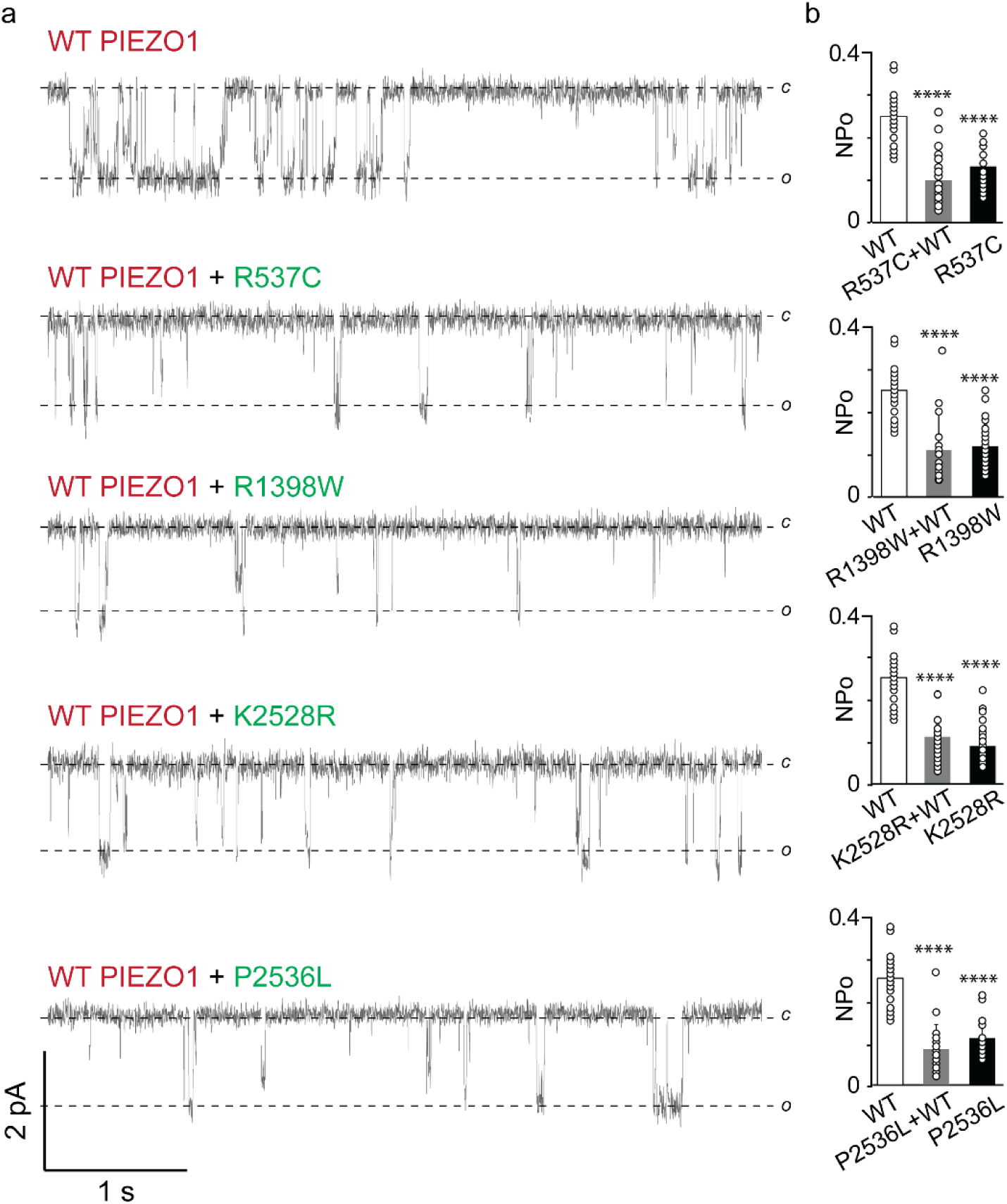
Functional dominant negative effect of OA-associated PIEZO1 mutations. **(a)** Single-channel current recordings of WT, mutants, and WT/mutant co-expressed PIEZO1 channels obtained from HEK293T^ΔP1^ cells. WT PIEZO1 is tagged with tdTomato and mutant PIEZO1 is tagged with GFP. Co-expression of tdTomato and GFP is used to identify single cells expressing both WT and mutant channels. Data acquired at -60 mV and -30 mm Hg pressure. **(b)** Comparison of steady state open probability between WT, mutants, and WT/mutant co-expressed PIEZO1 channels. ****p<0.0001 and c =closed, o =open, pA = picoamperes, NPo = normalized open probability. Experimental replica n between 16 and 27 individual patch recordings for each condition.

### Yoda1 Rescues Loss-of-Function OA-associated Variants

Since its discovery in 2015, Yoda1 – a specific chemical modulator of PIEZO1-has emerged as a valuable tool to study PIEZO1 by increasing PIEZO1 functional activity in different assays^50^. For example, in single channel electrophysiology recordings it increases the open probability of the channel, in whole cell recordings it delays inactivation, and in cell-based fluorescence assays it allows calcium flux via PIEZO1 channels in the absence of applied forces (pressure, tension etc)^41^. Here, we tested the rescue effect of Yoda1 on OA-associated loss-of-function PIEZO1 mutations by supplementing 20 µM Yoda1 in the patch pipette in cell-attached single channel recordings **(Fig 4).** As previously published, WT PIEZO1 responded to Yoda1 by exhibiting a 2-fold increase in open probability^41^. Interestingly, all the mutants responded to Yoda1 with fold increase in open probability as R537C = 1.5-fold, R1398W = 3-fold, K2528R = 4-fold, and P2536L = 2.5-fold (**Fig. 4b).** Additionally, no change in the single channel currents were observed with and without Yoda1, suggesting that Yoda1 only changes the gating properties of the mutant channels **(Fig. 4a).** In summary, the functional assays showed that Yoda1 increases the open probability of all the familial mutants when compared to WT **(Fig 4c).**

**Figure 4.**
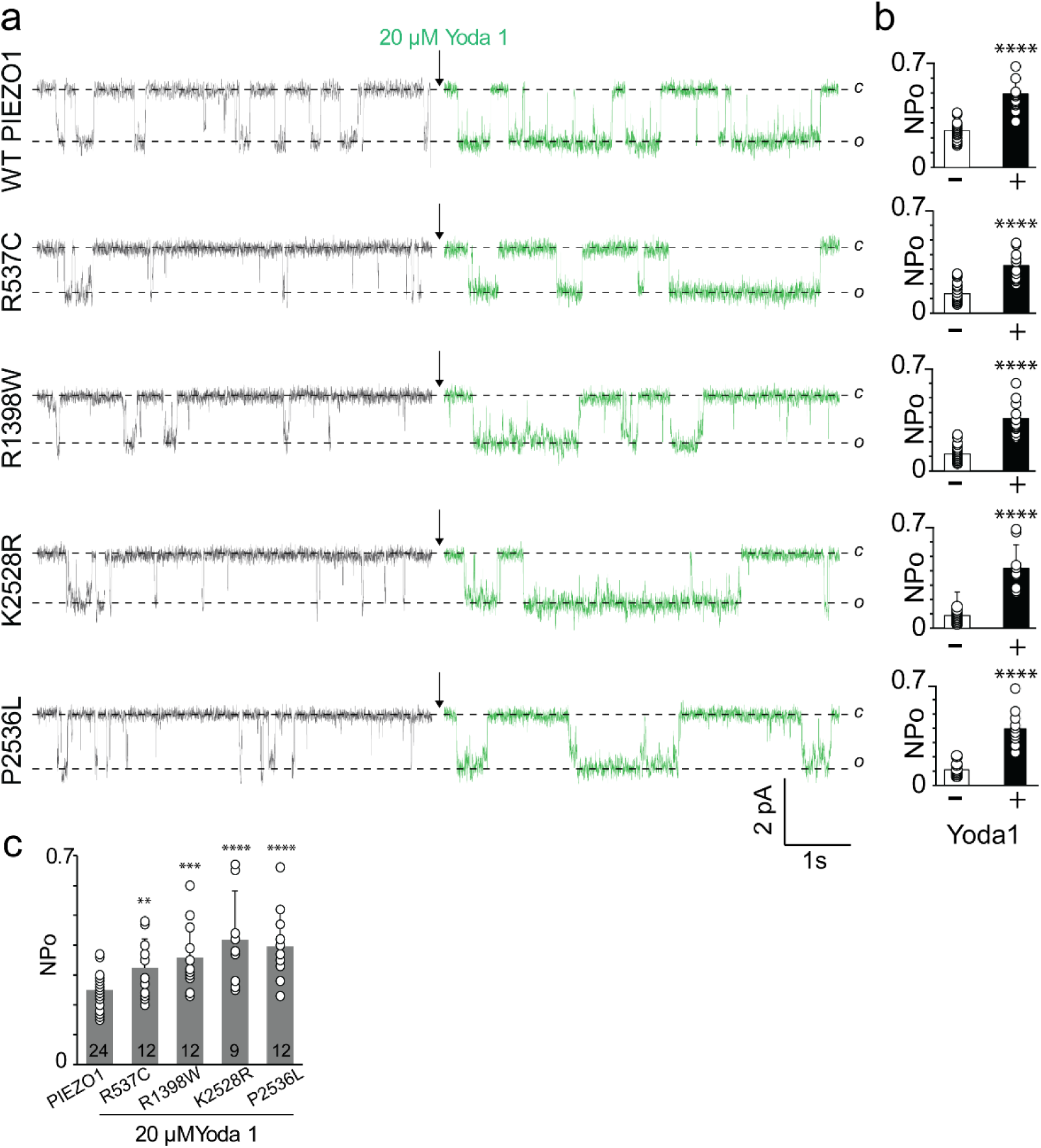
Chemical modulation of WT and OA-associated PIEZO1 channels by Yoda1. **(a)** Representative single-channel current recordings of WT and mutant PIEZO1 channels. The data was obtained from cell-attached patches at -60 mV and -30 mm Hg applied pressure. *o* and *c* denote open and closed state of the channel. **(b)** Steady state open probability of WT and mutant PIEZO1 channels with and without Yoda1 (n>9). **(c)** Comparison of steady state open probability of WT PIEZO1 channels in the absence of Yoda1 and mutant PIEZO1 channels in the presence of Yoda1. Experimental replica numbers are shown in the bar graph (n>9). **p<0.01, ***p<0.001, ****p<0.0001

In addition to single-channel activity, we evaluated whole cell calcium influx via cell-based fluorescence assay using Fluo-4 dye in HEK293S^ΔP1^. The cells were transfected with either WT PIEZO1 or familial mutants and pre-loaded with Fluo-4 dye and probenecid before treatment with 20µM Yoda-1 (solvent 0.2% DMSO) or 0.2% DMSO control **(Fig 5a).** In line with our previous data^41^, addition of Yoda1 affected intracellular calcium influx in cells expressing WT and the familial mutants, suggesting that channels are chemically stimulated in the absence of applied mechanical forces (such as pressure). Cells transfected with WT or mutant PIEZO1 when treated with DMSO did not induce calcium influx or did the mock transfection with Yoda1 **(Fig. 5).** Total cell fluorescence of R1398W and K2528R are not statistically different than WT PIEZO1, while R537C and P2536L achieved 65% and 80% of WT channel activity, respectively **(Fig. 5b),** indicating that the degree of calcium influx is dependent on the mutant expressed. In conclusion, using two functional assays, we show that the familial PIEZO1 variants’ gating properties can be restored by Yoda1.

**Figure 5.**
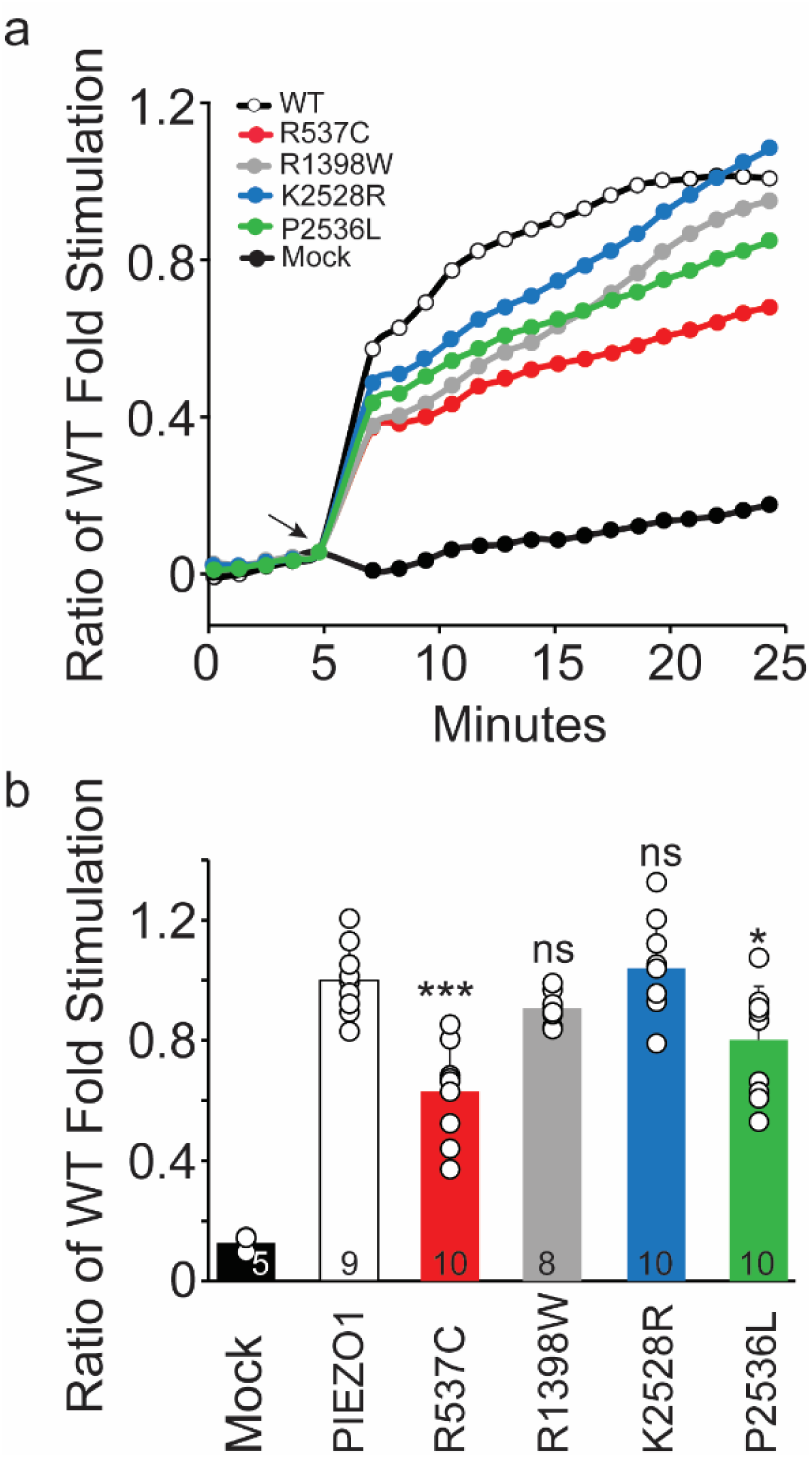
Yoda1 specifically stimulates PIEZO1-dependent calcium Influx in HEK293SΔP1 Cells expressing WT and OA-associated PIEZO1 mutants. **(a).** Fluorescent detection of intracellular calcium influx via Fluo-4 dye with or without Yoda1 in cells expressing WT or indicated mutant PIEZO1 channels. HEK293SΔP1 cells transfected with indicated constructs and pre-loaded with Fluo-4 dye before treatment with 20µM Yoda-1/0.2% DMSO or 0.2% DMSO control. Addition of Yoda1 or DMSO at 5min (black arrow). **(b)** Quantification of calcium influx indicated by normalized cell fluorescence induced by Yoda1. Data points reflect maximal activated signal prior to addition of ionophore. Individual transfections with R1398W and K2528R are not statistically different from WT (p>0.05). Transfection with R537C (P<0.0005) and P2536L(P<0.05) yielded decreased activity statistically significant than WT. Mock = empty vector transfection. Data point represent n>5 (indicated in bar graph) from individual wells.

## Discussion

Here we describe the association of four independent *PIEZO1* coding mutations with familial age-associated erosive hand OA (EHOA) or interphalangeal (IP) joint OA. Functional analyses of the familial OA-associated PIEZO1 proteins demonstrated that all four mutations are hypomorphic, since the mutations affect PIEZO1 function by reducing the pressure-induced open probability of the channel. In contrast with the familial mutations, we show at the molecular level for the first time that a previously identified GWAS *PIEZO1* mutation^32^ associated with reduced OA progression is hypermorphic as it increases the open probability of the channel. The most parsimonious interpretation of the human genetic data is that reduced PIEZO1 activity increases OA susceptibility while increased PIEZO1 activity reduces OA susceptibility, although there are potential confounding differences between the familial and GWAS cohorts. The families we studied have well defined EHOA or IP joint OA that occurs without acute or traumatic injury to the joint. The GWAS cohorts were identified based on knee and hip OA and injury status was not described and the GWAS study was based on whether individuals with OA progressed to need a joint replacement. Although the human *PIEZO1* GWAS allele (p.F2458L) was associated with a reduction in joint replacement, no alleles of *PIEZO1* have been associated with OA susceptibility previously^43,51–56^. Therefore, the role of p.F2458L on primary OA susceptibility remains unresolved and merits further in-depth studies.

Our genetic and functional analyses indicated that normal PIEZO1 activity is necessary to maintain joint homeostasis. Individuals with familial hypomorphic *PIEZO1* mutations develop EHOA or IP joint OA in the absence of injury or hyperphysiological load. Our data is in apparent contrast to previous *in vitro* studies indicating that PIEZO1 has a limited role in under normal loading conditions^15–19^. Based on our genetic and *in vitro* functional analyses, we propose that PIEZO1 has important roles in the normal response to loading in addition to the well characterized PIEZO1 response to acute injury or hyperphysiological loading. Hyperphysiological activation of PIEZO1 in cartilage induces inflammatory and catabolic gene expression, cell death, and senescence^3,15–19,22^, which supports the hypothesis that activation of PIEZO1 in the joint promotes development of OA. Recent transcriptomic data examining mechanical activation of PIEZO1 in chondrocytes is consistent with previously published data, supporting a central role of PIEZO1 in hyperphysiological load and promoting OA-associated gene expression^21^. The studies also discovered unexpected pathways regulated by chemical activation of PIEZO1. A two-week treatment of chondrocytes with the chemical agonist Yoda1, resulted in expression of anabolic and pro-chondrogenic pathways and an increase of sulfated glycosaminoglycans. In sum, our human genetic data and *in vitro* data indicate that PIEZO1 might have context dependent activity in homeostasis vs acute injury based. Several mutant mouse models of OA have context-specific effects, including the *Trpv4* mutant^57,58^, altering susceptibility to PTOA and age-dependent OA in different directions^59–61^.

A functional PIEZO channel assembles as a trimer. It is not yet proven (as per structural and biochemical analysis) whether PIEZO channels exist as hetero-trimers or if PIEZO1/2 hetero-trimer can form a functional channel. Although it has been reported that dominant hypomorphic alleles *PIEZO1* associated with left ventricular outflow tract obstructions act as dominant negative^62^. Our electrophysiology single channel data indicated that all four OA-associated familial PIEZO1 mutations function as dominant-negative channels, with a strong possibility of forming hetero-trimeric channels with WT subunit. This dominant-negative effect may be a common mechanism shared among dominant hypomorphic alleles of *PIEZO1*^62^, since no homozygous mutations are identified yet in any human disorders.

There are >25 mutations (both gain- and loss-of-function) in *PIEZO1* associated with various human diseases^5,63^, yet our studies are the first to show a clear association with increased OA susceptibility. We do not understand why individuals with the OA-associated familial mutations specifically develop OA. It is possible that chondrocytes (or other cells in the joint) are exquisitely sensitive to these mutations. Three of the mutations (P2536L, R1398W, and K2528R) cluster together on the intercellular side of the protein. These mutations may disrupt tissue specific protein-protein interactions necessary for normal PIEZO1 activity. We know that there are many genetic, physical and environmental risk factors that contribute to OA susceptibility^33,36,64^. There may be specific risk factors (e.g., obesity or exposure to environmental factors) shared among family members that in combination with *PIEZO1* alleles contribute to increased OA susceptibility. While we cannot exclude this possibility, it is unlikely given the *PIEZO1* alleles appear to be completely penetrant in our OA families. Finally, while our families do not have clear phenotypes associated with known loss-of-function *PIEZO1* alleles^41,49,62,65^, some affected individuals also have osteopenia, osteoporosis, several cardiac defects (atrial fibrillation and heart murmur), and hypertension. It is possible that the OA-associated mutations are contributing to these phenotypes in other organs.

The combined cellular, molecular, and genetic data suggest the role of PIEZO1 in joint homeostasis and OA may be more complex than previously appreciated. *Piezo1* null mice are embryonic lethal and the role of PIEZO1 in articular chondrocytes remain unclear^4,66^. Deletion of *Piezo1* and *Piezo2* (using *Gdf5-Cre*) in mouse cartilage has no measurable effect on injury induced OA^67^. In contrast, deletion of *Piezo1* (using *Col2a-Cre*) protected against injury induced OA^68^. It is difficult to determine the primary contribution of PIEZO1 to OA in these studies. The *Piezo1^Col2a1Cre^* mice have defects in endochondral ossification and other bone phenotypes, all of which may contribute to OA development independent of PIEZO1 function in mature articular chondrocytes^68^. The use of spatially restricted null alleles of *Piezo1* in the context of acute injury may not reflect the contribution of PIEZO1 in the whole joint during injury or age-associated OA. PIEZO1 may have context dependent and tissue specific roles in other tissues of the joint such as synovium, immune cells, or the infrapatellar fat pad, which would not be uncovered by used cartilage specific deletion of *Piezo1*^69,70^. Generation of mice with human-specific *PIEZO1* OA alleles will be important for determining how alteration of PIEZO1 channel activity contributes to disease onset and progression^34^.

In sum, we have discovered rare *PIEZO1* coding mutations associated with familial age-associated erosive hand OA (EHOA) or interphalangeal (IP) joint OA. Our functional analysis indicates that these mutations act as dominant-negative alleles to reduce the open probability of the channel. Furthermore, we demonstrate that the *PIEZO1* GWAS allele associated with reduced OA progression has increased channel activity. Given the genetic and functional data, we propose that PIEZO1 likely has context dependent effects in injury induced vs age-associated OA. The generation of mice with these human alleles will allow us to precisely define how modification of PIEZO1 activity contributes to OA susceptibly in different contexts.

## Methods

### Study approval

The Institutional Review Board of the University of Utah and the Resource for Genetic and Epidemiologic Research approved this study. Written informed consent was obtained under the guidance of the Institutional Review Board of the University of Utah.

### Diagnostic and procedure codes used to identify individuals with osteoarthritis

Our coding strategy used to identify individuals with EHOA and distal and proximal interphalangeal joint OA has been previously described^5,34,36^. Briefly, the following diagnostic codes (ICD-9 and ICD-10) and procedure codes (CPT - Current Procedural Terminology) were used to identify affected individuals. EHOA: ICD-10 M15.4 (Erosive (osteo)arthritis). Distal and proximal interphalangeal joint OA: CPT: 26862, 26863, 26860, 26861 (arthrodesis, interphalangeal joint, with or without internal fixation) and 26535, 26536 (arthroplasty, interphalangeal joint). ICD-9: 715.14 (osteoarthritis, localized, primary, hand). ICD-10: M19.04x (primary osteoarthritis, hand).

Individuals diagnosed with any of the following codes were excluded: ICD-9: 714.0 (rheumatoid arthritis), 714.2 and 714.3 (rheumatoid arthritis and other inflammatory polyarthropathies). ICD-10: M05.xxx (rheumatoid polyneuropathy with rheumatoid arthritis), M06.xx (other rheumatoid arthritis), or M08.xxx (juvenile arthritis). Individuals were asked if they were diagnosed with psoriatic arthritis, gout, or had a traumatic or acute injury to the affected joint. If they answered yes, they were excluded from the study.

### Whole exome sequencing and analysis

Families with a statistically significant increase in incidence of OA that appeared to segregate as a simple dominant Mendelian trait were identified and selected for whole exome sequencing (WES)^33,34^. WES and analysis was performed using genomic DNA isolated from whole blood or saliva as previously described^35^. Briefly, libraries were prepared using the Agilent SureSelect XT Human All Exon + UTR (v8) kit followed by Illumina NovaSeq 6000 150 cycle paired end sequencing. We followed best practices established by the Broad Institute GATK for variant discovery (https://gatk.broadinstitute.org/hc/en-us). Analysis of variants was performed with ANNOVAR (http://annovar.openbioinformatics.org/en/latest/)^71^ and pVAAST (http://www.hufflab.org/software/pvaast/)^72^ in concert with PHEVOR2 (http://weatherby.genetics.utah.edu/phevor2/index.html)^73^. pVAAST is a probabilistic search tool that classifies variants with respect to the likely effect they have on gene function. It incorporates information including position of a variant within a gene, cross-species phylogenetic conservation, biological function, and pedigree structure. PHEVOR2 works in concert with the output of pVAAST to integrate phenotype, gene function, and disease information for improved power to identify disease-causing alleles.

### PIEZO1 clone generation

The mouse PIEZO1-GFP fused gene was synthesized using GenScript’s Clone EZ service. OA point mutations were generated from this parent clone, also using GenScript’s Clone EZ. For calcium flux assay, a Myc tag was introduced following amino acid 2422 and GFP was deleted to avoid fluorescence signal from fused fluorophore. All nucleotide sequences were sequence verified after every maxi prep DNA isolation. Primers for sequencing were as follows:

R537C (forward): 5’-CTGGTGGTCCTGTCACTTTC -3’

R1398W (forward): 5’-GGGACTGCCTCATCCTCTATAA -3’

K2528R and P2536L and 2422-Myc tag for all mutants used the following primers:

(forward): 5’-TTCCCCATCTCTTCCCCAAG -3’

(reverse): 5’-GGAAGATGAGCTTGGCGTATAG -3’

### Cell culture and transient transfection

Method for calcium flux experiments is as previously published^41^. Briefly, adherent PIEZO1 (HEK293S^ΔP1^) knock-out cells were cultured to near confluency (80%) in 1xDulbecco’s modified Eagle’s medium (DMEM) supplemented with 10% fetal bovine serum (FBS; Sigma) and 1x GlutaMAX (Gibco), harvested with 0.05% trypsin/1xEDTA, and plated onto poly-D-lysine (Gibco)-coated 96-well black-sided/clear-bottom plates (Costar 3603) at sufficient density to achieve near-confluency after 24 hrs. Modified manufacturer’s protocols used to transiently transfect cells using Lipofectamine 3000 (ThermoFisher, Waltham, MA). Cells were incubated 24 hours at +37°C and 8% CO2 to allow for adherence and expression.

### Electrophysiology

Transfected HEK293T^ΔP1^ cells were visualized using Nikon eclipse Ti2 microscope and C11440 Orca-Flash 4.0 LT digital camera (Hamamatsu). Cell-attached recordings of pressure-activated currents in HEK293T^ΔP1^ cells expressing the WT PIEZO1, OA-associated familial and GWAS point mutations were performed using Axopatch 200B amplifier and Digidata 1550B digitizer (Molecular Devices). Currents were acquired with pClamp 10.7 software at a sampling frequency of 10 kHz. Recording patch pipettes of borosilicate glass (Sutter Instruments BF150-86-10) were pulled and fire-polished to a tip resistance of 4 to 6 MΩ. The bath solution contained (in mM): 140 KCl (Fisher Bioreagents, BP366-500), 10 HEPES (Gibco, 15630080), 1 MgCl2 (Millipore Sigma, 63069), 10 glucose (Sigma, G8270-1KG), pH 7.3 (pH adjusted with KOH (LabChem LC193702). The pipette solution contained (in mM): 130 NaCl (Fisher Brand, S271-3), 5 KCl, 10 HEPES, 10 TEA-Cl (Acros Organics, 150901000), 1 CaCl2 (Millipore Sigma, 21115), 1 MgCl2 pH 7.3 (pH adjusted with NaOH). When mentioned, 20 µM Yoda1 (Tocris, 5586/10) was supplemented in the pipette solution. The bath electrode was grounded such that at positive applied potential, cations moved from pipette to bath (inward currents). Mechanical stimulation was applied via recording pipette using a high-speed pressure clamp system (ALA Scientific Instruments). Single-channel events are shown as downward deflection (inward currents) acquired at -60 mV (to match conventional direction of ion flow). The data was analyzed using Clampfit 10.7. The Normalized Open Probability was calculated by: NPo = *A*o / *A*c + *A*o. A_O_ and Ac are the areas under the curve of open and closed components of all-point histograms respectively. The areas under the curve of histograms were calculated using Clampfit analysis program and via Gaussian fits to the data.

### Calcium flux assay

Fluo-4 AM NW dye and probenecid (Molecular Probes) were prepared according to manufacturer’s instructions with dye spiked to a final 4 mM CaCl_2_ concentration to enhance signal. Yoda-1 (Tocris) was prepared as 10 mM stock in 100% DMSO (Molecular Probes). HEK293S^ΔP1^ cells were seeded and reverse transfected in a 96-well plate and incubated 24 hours at +37°C and 8% CO_2_ before aspirating media and loading cells with dye/probenecid solution for 40 minutes at +37°C and 8% CO_2_. After loading cells with dye, baseline fluorescence was measured for 5 minutes followed by an 18-minute measurement of activation with 20 µM Yoda-1 (Tocris)/0.2% DMSO (Molecular Probes) or 0.2% DMSO alone. A23187 calcium ionophore was then added and cells assayed for 25 minutes to measure maximum cellular calcium signal. Calcium flux was assayed on a BioTek Synergy H1 at +37°C (Ex488/Em518) with 70 second time points.

### Statistics

All-point current amplitude histograms of the single-channel data (ranging from 15 to 30 s stretch) were constructed in Clampfit. The single-channel current values were extracted after fitting the Gaussian function to the data. Each mean value is an average of at least 9 or at most 27 individual patch recordings. 8-12 individual coverslips with transfected cells, either WT PIEZO1 or mutants, were used to acquire single channel data. The open probability of the channel was calculated exclusively from the records, where at least 10 s of data was recorded both before and after pressure application. The number of channels in the patch is determined by fitting a Gaussian curve to all-point current histograms to individual open peak. Group data (conductance bar graphs) are presented as Mean ± Standard Deviation where ∗ p < 0.05; ∗∗p < 0.01, ∗∗∗p < 0.001, ****p<0.0001, not significant (ns) p > 0.05. Unpaired two-tailed t-tests were used for comparison between the two groups.

## Data Availability

All data produced in the present study are available upon reasonable request to the authors

## Acknowledgments

We thank the families who participated in this study. This work was funded by the Skaggs Foundation for Research (MJJ), the Utah Genome Project (MJJ), the Arthritis National Research Foundation (MJJ – 707634), the National Institute on Aging (MJJ – R21AG063534-01A1), the National Institute of Arthritis and Musculoskeletal and Skin Diseases (MJJ and NHK – 1R01AR082973), and an LS Peery MD Medical Student Research Traineeship (DJM). RS is a W.W. Caruth, Jr. Scholar in Biomedical Research, supported by NIH grant (RS – R01GM142024) and UT Southwestern Medical Center Endowed Scholar Program.

## Author contributions

MJJ, NHK, and RS conceived and designed the study, collected and analyzed data, and wrote the manuscript. KAN collected and analyzed data and provided feedback on the manuscript. EN, DT, CM conducted experiments and analyzed data.

## Materials & Correspondence

Correspondence and material requests should be.

## Data availability

Data is available upon reasonable request. addressed to RS

